# A cross-sectional study of the neuropsychiatric phenotype of *CACNA1C*-related disorder

**DOI:** 10.1101/2022.06.22.22276784

**Authors:** Rebecca J. Levy, Katherine W. Timothy, Jack F. G. Underwood, Jeremy Hall, Jonathan A. Bernstein, Sergiu P. Paşca

**Affiliations:** Division of Medical Genetics in the Department of Pediatrics, Stanford University, Stanford, CA 94305, USA; Division of Child Neurology in the Department of Neurology, Stanford University, Stanford, CA 94305, USA; Timothy Syndrome Foundation, PO Box 1875, Ellicott City, MD, USA; Neuroscience & Mental Health Innovation Institute, Cardiff University, Wales, UK CR24 4HQ; Stanford Brain Organogenesis, Wu Tsai Neuroscience Institute, Stanford, CA 94305, USA; Department of Psychiatry and Behavioral Sciences, Stanford University, Stanford, CA 94305, USA

## Abstract

*CACNA1C* encodes the voltage gated L-type calcium channel CaV1.2. A specific gain of function pathogenic variant in *CACNA1C* causes Timothy syndrome type 1 (TS1) with cardiac long QT syndrome, syndactyly, and neuropsychiatric symptoms. Recent case series highlighted a broader spectrum of *CACNA1C*-related disorder (CRD) that includes isolated cardiac disease, isolated neurologic deficits, and TS, but it is unknown how the clinical presentation of other CRD variants relate to neural defects. We surveyed individuals with CRD to define the neuropsychiatric and developmental phenotype in an effort to guide future research into the role of calcium channels in neural development. Among 24 participants the most common neuropsychiatric symptoms and/or diagnoses were developmental delay in 92%, incoordination in 71%, hypotonia in 67%, autism spectrum disorder in 50% (autistic features in 92%), seizures in 38%, and attention deficit hyperactivity disorder in 21% of participants. There were no significant differences in symptoms between participants with and without long QT syndrome. These findings indicate the key role of CaV1.2 in brain development and the clinical importance of screening and therapeutically addressing neuropsychiatric symptoms in all individuals with CRD.

## Introduction

Voltage gated L-type calcium channels (LTCC) are critical for transforming chemical to electrical signals in the nervous system and other electrically excitable tissues.^1,2^ Common variants in *CACNA1C*, which encodes the α-subunit of the CaV1.2 LTCC, are among the most replicable associations in genome-wide association studies of autism spectrum disorder (ASD),^3^ bipolar disorder,^3–6^ depression,^3^ and schizophrenia.^3,6–8^ The dominant gain of function variant p.G406R in exon 8A of *CACNA1C* causes Timothy syndrome type 1 (TS1), a syndromic form of cardiac long QT interval syndrome (LQTS) with syndactyly, multi-system involvement, and a high prevalence of neurodevelopmental symptoms.^9^ In the original TS1 case series 80% of participants had ASD, thus TS1 has the highest prevalence of syndromic ASD.^9^

Heterozygous microdeletions including *CACNA1C* are associated with intellectual disability and are not known to be associated with cardiac or syndromic features, suggesting channel loss of function via reduced expression may also impact neurodevelopment.^10–12^ De novo variants in *CACNA1C*, along with other genes, were enriched in a population with developmental delay.^13^ Additional *CACNA1C* variants were recently identified in a broad spectrum of both cardiac and neurodevelopmental disorders including: typical and atypical syndromic TS,^9,14–19^ structural heart disease with LQTS,^20–22^ isolated long or short QTS,^23–26^ and isolated neurologic symptoms.^27–30^ This is now referred to as the *CACNA1C*-related disorder (CRD) spectrum that encompasses a range of clinical features caused by pathogenic variants predicted to both increase and decrease channel function.^31,32^

A better understanding of the epidemiology, clinical spectrum, and biological phenotype of highly penetrant monogenic disease models such as CRD would help unravel how calcium signaling affects neuropsychiatric disorders more broadly and how modulation of calcium pathways could be leveraged for therapeutic purposes. A major gap in the field is that neuropsychiatric symptoms are not well characterized in the CRD literature beyond the early TS1 cohort and very recent neurologic-only CRD cohort.^9,27^ Case reports of CRD-related arrhythmia report few if any neuro-psychiatric symptoms and do not evaluate a temporal relationship with cardiac arrest or other hypoxic risk factors. Individuals with CRD have never been systematically assessed for the prevalence of developmental, neurologic, and psychiatric symptoms, in part due to the rarity of this disorder. Moreover, there is neither clinical nor neuroscience research into whether treating channel dysfunction impacts neuropsychiatric outcomes.

## Results

### Demographics

Twenty-four survey participants with CRD had germline variants in *CACNA1C* (**Table 1**). One participant with mosaicism is not reported in this data although this individual reported symptoms that overlap with the cohort. Participants were from the United States and the United Kingdom. Unfortunately, three participants were deceased prior to study enrollment at a range of 1.6 to 6 years old. Mean age in the cohort at time of completion of survey was 9.8 years (range 0.4 years to 37 years) including the deceased participants and 10.6 years among surviving participants. Nine of the participants (37.5%) were female. Two participants were monozygotic twins.

**Table 1.** CRD cohort demographics and symptoms. * Indicates deceased participant. Abbreviations: CRD: *CACNA1C*-related disorder; LQTS: long QT syndrome; TS1: Timothy syndrome type 1; TS2: Timothy syndrome type 2.

| ID | Genotype | Phenotype | Hypoxic event | Developmental delay | Neurologic symptoms | Psychiatric diagnoses |
| --- | --- | --- | --- | --- | --- | --- |
| 1 | p.G406R exon 8A | TS1 | Y | Y | Y | Y |
| 2 | p.G406R exon 8A | TS1 | Y | Y | Y | N |
| 3 | p.G406R exon 8A | TS1 | Y | Y | Y | Y |
| 4 * | p.G406R exon 8A | TS1 | Y | Y | Y | Y |
| 5 * | p.G406R exon 8A | TS1 | Y | Y | Y | N |
| 6 | p.G406R exon 8A | TS1 | Y | Y | Y | Y |
| 7 | p.G406R exon 8A | TS1 | Y | Y | Y | Y |
| 8 | p.G406R exon 8 | TS2 | Y | Y | Y | N |
| 9 | p.G406R exon 8 | TS2 | Y | Y | Y | Y |
| 10 | p.V1363L | Other LQTS CRD | N | Y | Y | N |
| 11 | p.C1021R | Other LQTS CRD | N | Y | Y | N |
| 12 | p.G402S | Other LQTS CRD | Y | Y | Y | Y |
| 13 | p.E2062X | Non-LQTS CRD | Y | Y | Y | Y |
| 14 | p.V403M | Non-LQTS CRD | N | Y | Y | Y |
| 15 | p.A1521P | Non-LQTS CRD | N | Y | Y | Y |
| 16 | Deletion of exon 3 | Non-LQTS CRD | N | N | N | Y |
| 17 | p.L658P | Non-LQTS CRD | N | Y | Y | N |
| 18 | p.L207R | Non-LQTS CRD | N | Y | Y | Y |
| 19 | p.V1167A | Non-LQTS CRD | N | Y | Y | N |
| 20 * | p.V1363M | Non-LQTS CRD | N | Y | Y | N |
| 21 | p.G1173_Q1175del | Non-LQTS CRD | N | Y | Y | Y |
| 22 | p.R1332Q | Non-LQTS CRD | Y | Y | N | Y |
| 23 | p.R1332Q | Non-LQTS CRD | N | Y | N | Y |
| 24 | p.T941I | Non-LQTS CRD | N | Y | Y | N |

We investigated genotype-phenotype correlation by considering the following sub-populations of CRD. We grouped participants by genotype and presence of LQTS at baseline, which is a well-defined major clinical criterion for TS. Seven participants have TS1 (defined as p.G406R in exon 8A), two have TS type 2 (TS2, defined as p.G406R in exon 8), three have other LQTS CRD (defined as a variant outside of p.G406R plus LQTS at baseline), and 12 have non-LQTS CRD (defined as a variant outside the p.G406R locus and absence of LQTS at baseline). Given the small number of TS2 and other LQTS CRD participants, analyses were performed as a comparison between all LQTS CRD (TS1, TS2, and other LQTS CRD) and non-LQTS CRD participants.

### Developmental risk factors

Overall, 54.2% of participants reported a major hypoxic event, defined as perinatal respiratory distress, perinatal hypoxic ischemic encephalopathy, or cardiac arrest (**Table 2**). Major hypoxic events tended to be more common in LQTS CRD than non-LQTS CRD participants (83.3% vs. 25%, Fisher’s exact test P = 0.012, Bonferroni corrected for α = 0.00167 for developmental comparisons).

**Table 2.** Percent of CRD participants with commonly reported neuropsychiatric symptoms. Percent of participants with reported neurologic and psychiatric symptoms for all participants and genetic subsets. Fisher’s exact test two tailed P-value was calculated on the number of participants with and without cardiac arrhythmia. Abbreviations: ADHD: attention deficit hyperactivity disorder; ASD: autism spectrum disorder; CRD *CACNA1C*-related disorder; dx: diagnosis (indicates report of a formal diagnosis); LQTS: long QT syndrome.

| Symptom (%) | All CRD (n = 24) | LQTS CRD (n = 12) | Non-LQTS CRD (n = 12) | Fisher's exact P-value |
| --- | --- | --- | --- | --- |
| Hypoxic event | 54.2 | 83.3 | 25 | 0.01 |
| Learning or intellectual disability | 70.8 | 66.7 | 75 | 1 |
| Incoordination | 70.8 | 66.7 | 75 | 1 |
| Dysphagia | 62.5 | 58.3 | 66.7 | 1 |
| Hypotonia | 66.7 | 50 | 83.3 | 0.19 |
| Seizures, epilepsy | 39.1 | 41.7 | 36.4 | 1 |
| Any psychiatric dx | 62.5 | 58.3 | 66.7 | 1 |
| ASD dx | 50 | 50 | 50 | 1 |
| Aggression, self-injurious behavior | 29.2 | 33.3 | 25 | 1 |
| ADHD dx | 20.8 | 16.7 | 25 | 1 |
| Anxiety dx | 16.7 | 16.7 | 16.7 | 1 |

Prematurity (birth before 36 weeks gestational age) was reported in 29.2% of all participants. Premature birth was not significantly more common in LQTS CRD than non-LQTS CRD participants (33.3% vs. 25%, Fisher’s exact test P = 1).

### Developmental phenotype

Developmental delay was present in 91.7% of all participants, with 91.7% of participants reporting motor delay and 75% reporting language delay (**Table 3**). Seventeen percent of participants were not yet able to walk and/or speak and other language and motor milestones were similarly affected. There was a trend that non-LQTS CRD participants were less likely to be able to learn to ride a bicycle, but there were no significant differences between participants with and without LQTS in the ability to achieve any of the milestones after multiple comparison correction.

**Table 3.** Percent of participants with delays and inability to achieve developmental milestones. The percent of all participants and subsets with and without cardiac arrhythmia are shown for presence of language and motor delay and then inability to achieve these developmental mile-stones. Bonferroni multiple comparison correction P <0.00167 for Supplemental Tabbles 1 and 2. Fisher’s exact test two tailed P-value was calculated on the number of participants with and without cardiac arrhythmia. Abbreviations: CRD: *CACNA1C*-related disorder; LQTS: long QT syndrome; TS1: Timothy syndrome type 1; TS2: Timothy syndrome type 2.

| Milestone (%) | All CRD<br>(n = 24) | LQTS CRD<br>(n = 12) | Non-LQTS CRD<br>(n = 12) | Fisher's exact P-value |
| --- | --- | --- | --- | --- |
| Language delay | 75 | 66.7 | 83.3 | 0.64 |
| Motor delay | 91.7 | 91.7 | 91.7 | 1 |
| Rolling | 8.3 | 8.3 | 8.3 | 1 |
| Sitting | 8.3 | 8.3 | 8.3 | 1 |
| Walking | 16.7 | 16.7 | 16.7 | 1 |
| Biking | 41.7 | 16.7 | 66.7 | 0.04 |
| Pincer | 8.3 | 8.3 | 8.3 | 1 |
| Utensils | 20.8 | 8.3 | 33.3 | 0.32 |
| First word | 16.7 | 8.3 | 25 | 0.59 |
| Speak in phrases | 20.8 | 8.3 | 33.3 | 0.32 |
| Follow a command | 16.7 | 8.3 | 25 | 0.59 |
| Smile | 4.2 | 8.3 | 0 | 1 |
| Imaginative play | 25 | 8.3 | 41.7 | 0.16 |
| Toilet trained day | 29.2 | 25.0 | 33.3 | 1 |
| Toilet trained night | 33.3 | 33.3 | 33.3 | 1 |

Among participants who achieved developmental milestones, the average age was higher than typical development across motor, language, and social domains and across all subtypes of CRD (**Figure 1A, B**). Participants with non-LQTS CRD tended to be more delayed on development of first spoken word and independent sitting compared to participants with LQTS CRD (**Table 4**).

**Figure 1.**
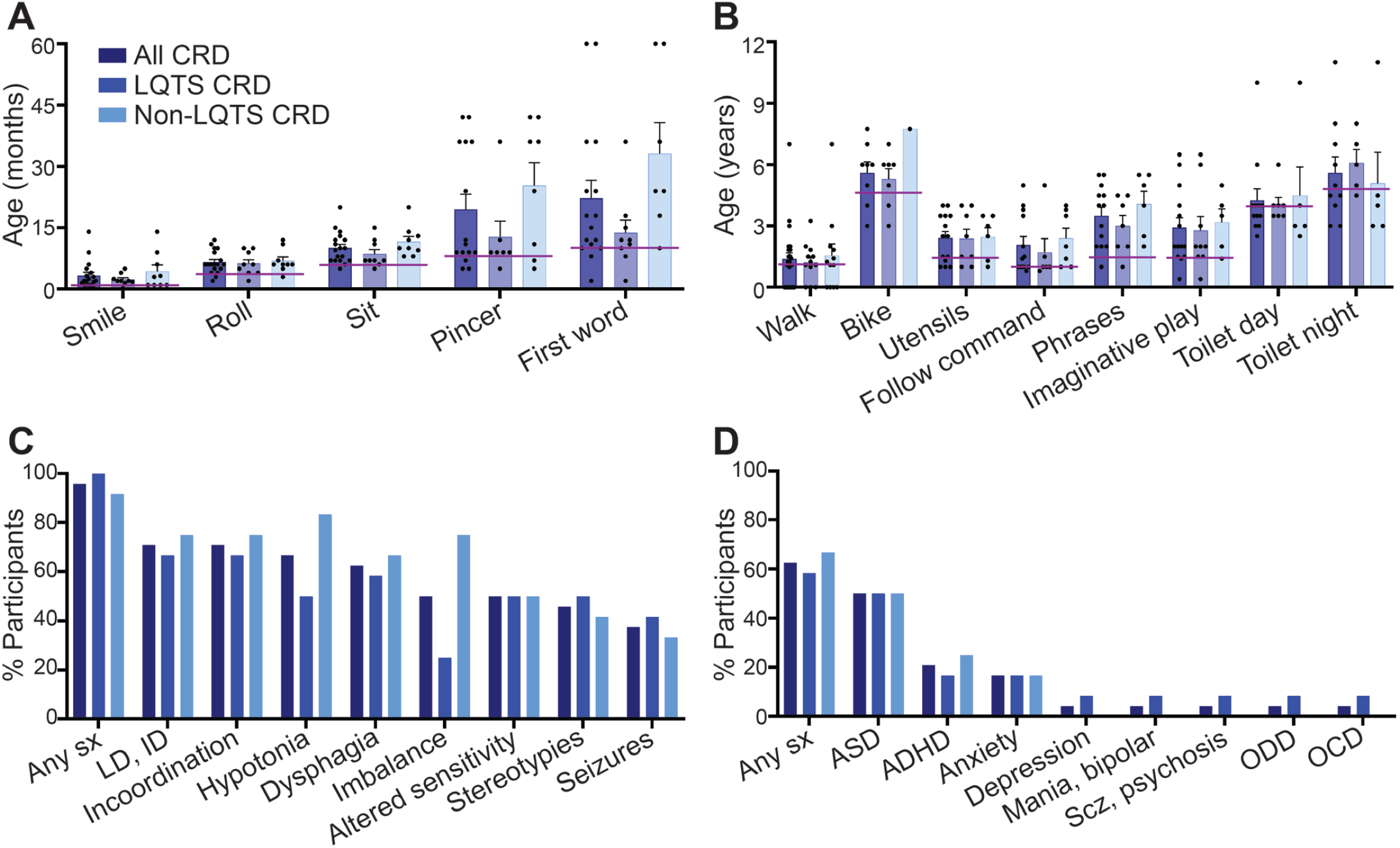
Developmental, neurologic, and psychiatric symptoms are highly prevalent in CRD but do not differ by history of LQTS. (**A**) Mean age in months or (**B**) years at which participants achieved developmental milestones. All participants are depicted next to participants with and without LQTS. There were no significant differences between subgroups. Age at achievement in 50% of typical population denoted by purple lines derived from the Denver II Developmental Scale.^42^ Error bars show standard error of the mean. (**C**) Percent of participants who reported the most common neurologic symptoms. There were no significant differences between subgroups. (**D**) Percent of participants who reported the most common psychiatric diagnoses. There were no significant differences between subgroups. Abbreviations: ADHD: attention deficit hyperactivity disorder; ASD: autism spectrum disorder; CRD: *CACNA1C*-related disorder: dx: diagnosis; ID: intellectual disability; LD: learning disability; LQTS: long QT syndrome; OCD: obsessive compulsive disorder; ODD: oppositional defiant disorder; Scz: schizophrenia; sx: symptoms.

**Table 4.**
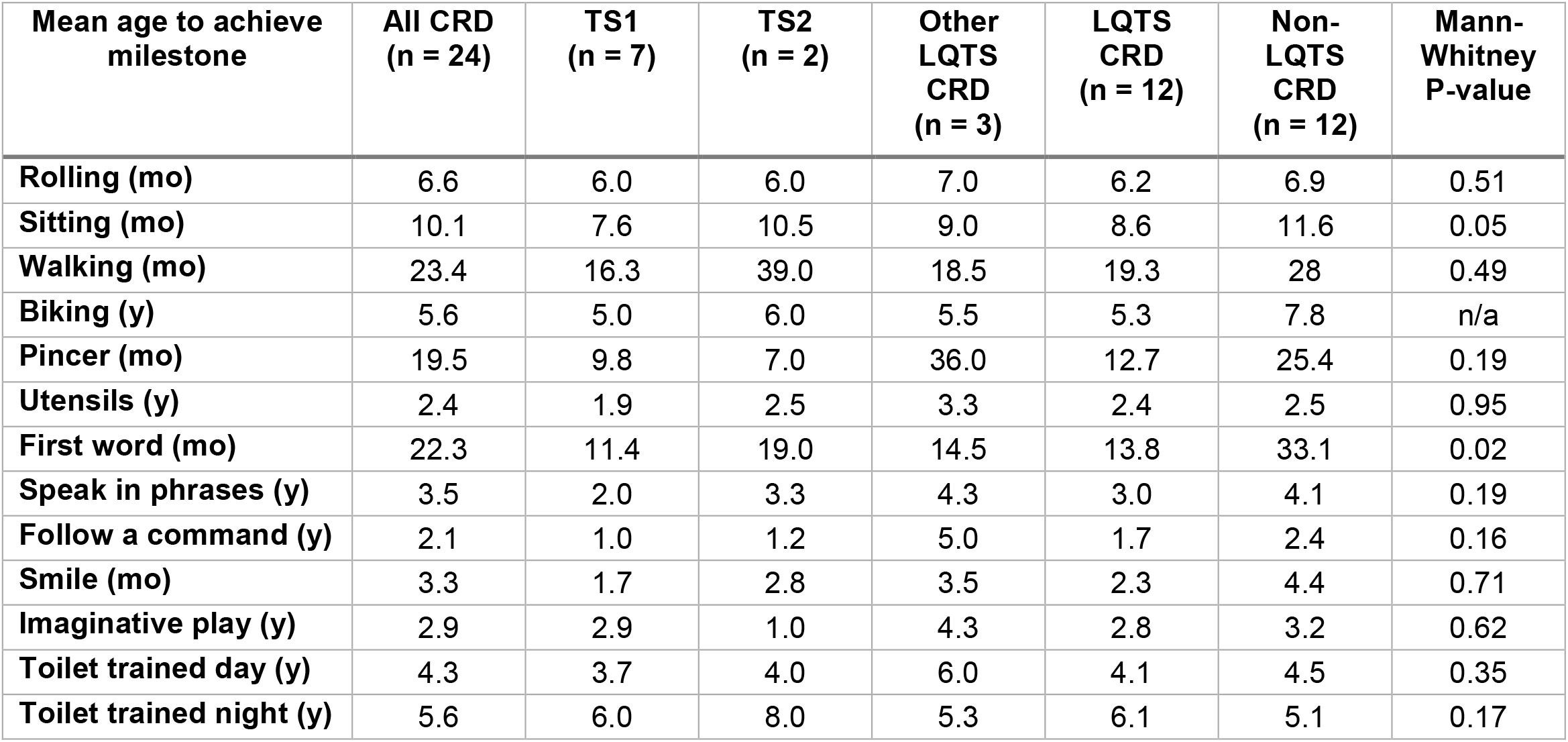
Mean age at which participants achieved developmental milestones. The Mean age at which participants achieved developmental milestones for all participants and subsets with and without cardiac arrhythmia. Students unpaired two tailed t-test was calculated on the mean age of participants with and without cardiac arrhythmia and p-values reported. Bonferroni multiple comparison correction P <0.00167 for Supplemental Tabbles 1 and 2. Abbreviations: CRD: *CACNA1C*-related disorder; LQTS: long QT syndrome; mo: months; n/a: not applicable, no value can be calculated due to sample size; TS1: Timothy syndrome type 1; TS2: Timothy syndrome type 2; y: years.

### Neurologic phenotype

Neurologic symptoms of any kind were reported in 95.8% of all participants (**Table 5**). The most common symptoms included learning disability and/or intellectual disability, incoordination, dysphagia, hypotonia, and altered sensitivity to noxious stimuli (**Figure 1C**). Seizures or epilepsy were reported in 37.5% of all participants. Of note, while intellectual and learning disabilities were common, 33.3% of participants reported highly accurate memory or other above-average skills for age. No symptoms were significantly different between participants with or without arrhythmia. There was a trend that imbalance and abnormal extraocular movements were more common in non-LQTS CRD.

**Table 5.** Percent of participants with reported neurologic symptoms. Percent of participants with reported neurologic symptoms for all participants and genetic subsets. Fisher’s exact test two tailed P-value was calculated on the number of participants with and without cardiac arrhythmia. Bonferroni multiple comparison correction P <0.0019. Abbreviations: CRD: *CACNA1C*-related disorder; LQTS: long QT syndrome; TS1: Timothy syndrome type 1; TS2: Timothy syndrome type 2.

| Symptom (%) | All CRD<br>(n = 24) | TS1<br>(n = 7) | TS2<br>(n = 2) | Other<br>LQTS<br>CRD<br>(n = 3) | LQTS<br>CRD<br>(n = 12) | Non-LQTS<br>CRD<br>(n = 12) | Fisher's<br>exact<br>P-value |
| --- | --- | --- | --- | --- | --- | --- | --- |
| **Any neurologic symptoms** | 95.8 | 100 | 100 | 100 | 100 | 91.7 | 1 |
| **Seizures or epilepsy** | 37.5 | 28.6 | 50 | 66.7 | 41.7 | 33.3 | 1 |
| **Very accurate memory** | 33.3 | 28.6 | 0 | 66.7 | 33.3 | 33.3 | 1 |
| **Language delay** | 75 | 57.1 | 50 | 100 | 66.7 | 83.3 | 0.64 |
| **Language production disorder** | 50 | 28.6 | 50 | 66.7 | 41.7 | 58.3 | 0.68 |
| **Receptive language disability** | 33.3 | 14.3 | 0 | 33.3 | 16.7 | 50 | 0.19 |
| **Learning or intellectual disability** | 70.8 | 57.1 | 50 | 100 | 66.7 | 75 | 1 |
| **Motor delay** | 91.7 | 85.7 | 100 | 100 | 91.7 | 91.7 | 1 |
| **Incoordination** | 70.8 | 57.1 | 50 | 100 | 66.7 | 75 | 1 |
| **Weakness** | 50 | 28.6 | 0 | 66.7 | 33.3 | 66.7 | 0.22 |
| **Hypotonia** | 66.7 | 28.6 | 100 | 66.7 | 50 | 83.3 | 0.19 |
| **Hypertonia, spasticity** | 20.8 | 14.3 | 0 | 33.3 | 16.7 | 25 | 1 |
| **Imbalance** | 50 | 14.3 | 0 | 66.7 | 25 | 75 | 0.04 |
| **Tremors** | 8.3 | 14.3 | 0 | 0 | 8.3 | 8.3 | 1 |
| **Muscle cramps** | 8.3 | 14.3 | 0 | 0 | 8.3 | 8.3 | 1 |
| **Contractures** | 4.2 | 0 | 0 | 33.3 | 8.3 | 0 | 1 |
| **Dysphagia** | 62.5 | 71.4 | 50 | 33.3 | 58.3 | 66.7 | 1 |
| **Abnormal extraocular movements, nystagmus** | 16.7 | 0 | 0 | 0 | 0 | 33.3 | 0.09 |
| **Stereotypies or repetitive movements** | 45.8 | 28.6 | 50 | 100 | 50 | 41.7 | 1 |
| **Echolalia or repetitive vocalizations** | 50 | 42.9 | 50 | 66.7 | 50 | 50 | 1 |
| **Tics** | 29.2 | 57.1 | 0 | 33.3 | 41.7 | 16.7 | 0.37 |
| **Headache** | 16.7 | 28.6 | 0 | 0 | 16.7 | 16.7 | 1 |
| **Sensory loss** | 8.3 | 14.3 | 0 | 0 | 8.3 | 8.3 | 1 |
| **Altered sensitivity to pain and/or temperature** | 50 | 42.9 | 50 | 66.7 | 50 | 50 | 1 |
| **Vision deficit** | 45.8 | 28.6 | 50 | 66.7 | 41.7 | 50 | 1 |
| **Hearing deficit** | 12.5 | 14.3 | 50 | 33.3 | 25 | 0 | 0.22 |
| **Poor sleep quality** | 50 | 71.4 | 50 | 66.7 | 66.7 | 33.3 | 0.22 |

### Psychiatric phenotype

A spectrum of psychiatric symptoms and diagnoses were reported. Sixty-three percent of participants reported a formal diagnosis of at least one psychiatric disorder (**Table 6**). The most commonly reported diagnoses were ASD in 50% of all participants, attention deficit hyperactivity disorder (ADHD) in 20.8%, and anxiety disorder in 16.7% (**Figure 1D**). When asked about symptoms, 91.7% of participants reported at least one behavioral feature associated with ASD, 54.2% reported a shorter attention span, 29.2% reported aggressive behavior towards self or others, and 25% reported a specific phobia. There were no significant differences in self-reported symptoms nor in reported formal diagnoses between participants with and without LQTS.

**Table 6.** Percent of participants with reported psychiatric symptoms and diagnoses. Percent of participants who reported experiencing psychiatric symptoms and who reported a formal diagnosis for all participants and genetic subsets. Fisher’s exact test two tailed P-value was calculated on the number of participants with and without cardiac arrhythmia. Bonferroni multiple comparison correction P <0.0029. Abbreviations: ADHD: attention deficit hyperactivity disorder; CRD: *CACNA1C*-related disorder; dx: diagnosis (indicates report of a formal diagnosis); LQTS: long QT syndrome; TS1: Timothy syndrome type 1; TS2: Timothy syndrome type 2.

| Symptom (%) | All CRD<br>(n = 24) | TS1<br>(n = 7) | TS2<br>(n = 2) | Other<br>LQTS<br>CRD<br>(n = 3) | LQTS<br>CRD<br>(n = 12) | Non-LQTS<br>CRD<br>(n = 12) | Fisher's<br>exact<br>P-value |
| --- | --- | --- | --- | --- | --- | --- | --- |
| Any psychiatric dx | 62.5 | 71.4 | 50 | 33.3 | 58.3 | 66.7 | 1 |
| Any autistic symptoms | 91.7 | 85.7 | 100 | 100 | 91.7 | 91.7 | 1 |
| Autism spectrum disorder dx | 50 | 57.1 | 50 | 33.3 | 50 | 50 | 1 |
| Short attention span | 54.2 | 57.1 | 50 | 66.7 | 58.3 | 50 | 1 |
| Hyperactivity | 20.8 | 28.6 | 0 | 0 | 16.7 | 25 | 1 |
| ADHD dx | 20.8 | 28.6 | 0 | 0 | 16.7 | 25 | 1 |
| Aggression, self-injurious behavior | 29.2 | 57.1 | 0 | 0 | 33.3 | 25 | 1 |
| Specific phobias | 25 | 28.6 | 0 | 33.3 | 25 | 25 | 1 |
| Frequent worrying | 16.7 | 28.6 | 0 | 33.3 | 25 | 8.3 | 0.59 |
| Anxiety dx | 16.7 | 28.6 | 0 | 0 | 16.7 | 16.7 | 1 |
| Social anxiety, agoraphobia | 12.5 | 14.3 | 0 | 33.3 | 16.7 | 8.3 | 1 |
| Frequent sadness or depressed mood | 4.2 | 14.3 | 0 | 0 | 8.3 | 0 | 1 |
| Depression dx | 4.2 | 14.3 | 0 | 0 | 8.3 | 0 | 1 |
| Manic symptoms | 4.2 | 0 | 0 | 0 | 0 | 8.3 | 1 |
| Mania or bipolar dx | 4.2 | 14.3 | 0 | 0 | 8.3 | 0 | 1 |
| Schizophrenia, psychosis dx | 4.2 | 14.3 | 0 | 0 | 8.3 | 0 | 1 |
| Oppositional defiant disorder dx | 4.2 | 14.3 | 0 | 0 | 8.3 | 0 | 1 |
| Obsessive compulsive disorder dx | 4.2 | 14.3 | 0 | 0 | 8.3 | 0 | 1 |

Of note, one older participant with TS1 was the only individual with diagnoses of obsessive-compulsive disorder, oppositional defiant disorder, depression, mania and/or bipolar disorder, and schizophrenia, in addition to ASD and anxiety disorder.

## Discussion

*CACNA1C*-related disorder is a syndromic cause of cardiac arrhythmia that also impacts neurodevelopment. We surveyed a cohort of 24 participants with CRD (out of approximately 200 individuals in the literature, global incidence unknown) to catalog more broadly what types of developmental, neurologic, and psychiatric symptoms they experienced and found a high prevalence of developmental delay, neurologic, and psychiatric symptoms.

The most commonly reported psychiatric symptoms were features of ASD in 92% of participants with a formal diagnosis of ASD in 50%. Other commonly reported symptoms included short attention span, aggressive behavior, and specific phobias (never before reported in this disorder). Of note, while 63% of participants reported at least one formal psychiatric diagnosis, many reported symptoms but lacked a formal diagnosis, raising concern that these individuals may not have received psychiatric clinical evaluation or support for symptom management.

Ninety-two percent of participants reported motor and/or language developmental delay and 96% reported at least one neurologic symptom. Some of the most common neurologic symptoms reported were learning or intellectual disability, incoordination, concerns for feeding safety, hypotonia, and altered sensory sensitivity. Seizures or epilepsy were reported in 38% suggesting that education about seizure semiology and safety could be beneficial for individuals with CRD in addition to detailed screening.

There are no significant differences in neuropsychiatric symptoms between participants with and without LQTS despite a trend toward a difference in the history of major hypoxic events. This suggests that the neuronal calcium channel alteration is the primary etiology for CRD neuropsychiatric symptoms and that cardiac-related hypoxic injury is a secondary contributing factor. These findings highlight the essential role of CaV1.2 in neurodevelopment.

Our data align with and reinforce findings from the contemporaneously conducted Rodan *et al*. study examining neurologic-only CRD.^27^ We found comparable prevalence of neuropsychiatric symptoms even in the absence of cardiac symptoms. Together, these results demonstrate that *CACNA1C* plays an important role in neurodevelopment, cognition, and behavioral control. Our cohort is gene-centric, not symptom-centric, permitting a less biased ascertainment of the prevalence of neuropsychiatric symptoms. Thus, our data help confirm prior literature on the presence and spectrum of neuropsychiatric and developmental disorders across all subtypes of CRD.^9,27–30^

Two prior publications reported on the same individual with atypical TS (with LQTS) and bipolar disorder who experienced episodic psychosis that improved on verapamil, as well as subjective improvement in cognition on this medication.^33,34^ We report one older participant who received multiple psychiatric diagnoses. These results suggest a need for longitudinal follow up to determine if other individuals with CRD develop bipolar disorder, OCD, and/or psychosis.

There are notable recent advances from animal and human models of CRD.^35^ Heterozygous *Cacna1c* knock out rats had altered associative learning,^36^ as well as alterations in network oscillations and hippocampal plasticity markers, which were rescued by the neurotrophin receptor agonist LM22B-10.^37^ A knock-in mouse model of TS2 displayed restricted and repetitive behaviors.^38^ In parallel, 2D neurons and cortical organoids derived from individuals with TS1 had altered gene expression, increased neurotransmitters (norepinephrine and dopamine), and aberrant differentiation.^39^ In forebrain assembloids, TS1-derived interneurons demonstrated abnormal migration that was related to increased myosin light chain phosphorylation and GABA-A receptor sensitivity.^40,41^ It remains to be determined how these cellular level abnormalities can cause network level alterations that manifest as developmental and psychiatric symptoms seen in rodent models and individuals with CRD. We hope that continued research from clinical studies partnered with basic neuroscience will hopefully yield answers about the role of *CACNA1C* in typical development, the pathophysiology of CRD, and insight into treatments.

### Limitations

We acknowledge that our study has limitations stemming from cohort size and survey-based data acquisition methods. Our sample size was inherently limited by participant availability since CRD is a rare genetic disorder, which meant we were only powered to detect a large difference between subgroups. This also prevented a confirmatory replication study and more comprehensive statistical sub-analyses such as comparisons by hypoxic exposure and genotype-phenotype correlations. We were further limited to participants with sufficient English fluency to complete the survey. We classified participants by self-reported LQTS at baseline when there is a spectrum of QT interval involvement that may present later in life or under exercise stress.

This study was designed as a preliminary evaluation to determine if the subjective observations by caregivers and medical professionals that individuals with CRD often had developmental and neuropsychiatric symptoms was in fact correct. This study therefore involves a participant or caregiver-report based survey which, while based on neurodevelopmental and psychiatric symptom inventories, does not incorporate validated diagnostic tools. We recognize that this limits the accuracy of the data. Moreover, a survey approach may incorporate several forms of bias such as selection/ascertainment, recall, self-reporting, and/or diagnostic access biases. We are addressing some of these limitations via an ongoing deeper phenotyping study incorporating standardized assessment measures within this cohort.

## Conclusions

In a self- or caregiver-reported survey of 24 individuals with *CACNA1C*-related disorder we found pervasive developmental, neurologic, and psychiatric symptoms, including a 50% prevalence of ASD. These data are in alignment with prior publications on the risk of ASD and neurodevelopmental disorders in CRD and expand the spectrum of symptoms in this disorder. There were no significant differences in symptom prevalence between participants with and without long QT syndrome, suggesting that there are more similarities than distinctions when it comes to the impact of *CACNA1C* variants on the brain. In the absence of genotype-phenotype correlation, individuals with CRD should be screened and monitored for a broad spectrum of neuropsychiatric symptoms throughout their lifespan in order to properly diagnose, treat, and support maximal development. These findings will also inform research into the role of *CACNA1C* and calcium channels in neurodevelopment and neural function.

## Data Availability

Most of the data generated and analyzed during this study are available in the tables and supplemental files. Full data are available from the corresponding author on reasonable request to protect the individual privacy of the participants.

## Acknowledgements

We acknowledge the participating families as well as the Timothy Syndrome Alliance (timothysyndrome.org) and the Timothy Syndrome Foundation (timothysyndromefoundation.org). Dr. Victor Ritter from the Stanford Quantitative Sciences Unit reviewed and advised on statistical analyses. This paper was typeset with the bioRxiv word template by @Chrelli: www.github.com/chrelli/bioRxiv-word-template

## Author contributions

RJL, KWT, JFGU, JH, JAB, and SPP conceived and designed the study. RJL, KWT, and JAB generated the survey. KWT, RJL, JFGU, JH, and SPP recruited participants and RJL consented all participants and analyzed the data. All authors read and approved the final manuscript.

## Materials and Methods

### Data collection

Participants were enrolled in the ongoing study of neuropsychiatric and developmental symptoms in CRD, which was approved by the Stanford University Institutional Review Board. Participants were identified via the Timothy Syndrome Foundation, the Timothy Syndrome Alliance, and communication with medical providers. Legally authorized representatives gave written informed consent for their children and/or dependents. Data collection was via a secure online survey hosted in RedCap. Survey questions covered genetic variants, pre- and perinatal comorbidities that influence neurodevelopment, cardiac and medical complications, developmental milestones, neurologic symptoms, psychiatric symptoms, and medications. The survey questions are available upon request. Medical records were obtained from caregivers as available and included verification of *CACNA1C* variants for all participants.

### Statistics

Descriptive statistical analysis included percentages for categorical variables and mean and range for continuous variables. The multiple Mann-Whitney tests were used for comparison of continuous values and Fisher’s exact test for comparison of categorical variables between participants with and without cardiac involvement. Bonferroni correction for α was computed for each of the developmental, neurologic, and psychiatric comparisons. With our cohort size we had 80% power to detect a difference of 37% between the groups as a two-sided χ2 test with 5% significance with presumed 15% prevalence.

### Funding

An institutional grant from the Stanford Autism Working Group helped fund sample and data acquisition. JFGU is funded by a Wellcome Trust GW4-CAT Clinical Doctoral Fellowship (222849/Z/21/Z). JH is funded by MRC grant no. MR/R011397/1. SPP is a New York Stem Cell Foundation (NYSCF) Robertson Stem Cell Investigator, a Chan Zuckerberg Initiative (CZI) Ben Barres Investigator and a CZ BioHub Investigator. The Stanford REDCap platform (http://redcap.stanford.edu) is developed and operated by Stanford Medicine Research IT team. The REDCap platform services at Stanford are subsidized by a) Stanford School of Medicine Research Office, and b) the National Center for Research Resources and the National Center for Advancing Translational Sciences, National Institutes of Health, through grant UL1 TR001085. No funding body impacted design, analysis, or interpretation of this study.

